# Hypoxia-induced stromal and immune remodeling in gastric carcinoma: correlation of Hypoxia-inducible factor 1-alpha (HIF-1α) expression with cancer-associated fibroblast (CAF) subtypes and Programmed death-ligand 1 (PD-L1) expression

**DOI:** 10.64898/2026.08.10.26360060

**Authors:** Gazi Abdus Sadique, Md. Shahriar Mamun, Shantanu Biswas, Tamanna Afroz, Paromita Ghosh, Md. Washikur Rahman, Tanshina Afrin

## Abstract

**Background:** Gastric carcinoma remains a major cause of cancer-related mortality worldwide, with tumor progression increasingly recognized as a consequence of complex interactions within the tumor microenvironment. Hypoxia-induced signaling, cancer-associated fibroblast (CAF) heterogeneity, and immune checkpoint activation play critical roles in tumor progression and immune evasion. However, their integrated relationship in gastric carcinoma remains insufficiently characterized.

**Objectives:** To evaluate the expression of Hypoxia-inducible factor 1-alpha and its association with cancer-associated fibroblast subtypes and Programmed death-ligand 1 expression in gastric carcinoma.

**Methods:** This cross-sectional analytical study included 100 histologically confirmed gastric carcinoma cases from Satkhira Medical College. Immunohistochemistry was performed for HIF-1α, α-smooth muscle actin (α-SMA), interleukin-6/fibroblast activation protein (IL-6/FAP), and PD-L1. CAFs were subclassified into myofibroblastic CAFs (myCAFs) and inflammatory CAFs (iCAFs). Associations between biomarkers and clinicopathological variables were analyzed using chi-square test, Spearman correlation, and multivariate logistic regression. Receiver operating characteristic (ROC) curve analysis was used to assess model performance.

**Result:** High HIF-1α expression was observed in 55% of cases and demonstrated significant association with poor differentiation (p = 0.001), advanced tumor stage (p = 0.002), and lymph node metastasis (p = 0.001). iCAF predominance was significantly associated with poor differentiation (p = 0.003), advanced stage (p = 0.004), and nodal metastasis (p = 0.004). High PD-L1 expression was significantly associated with poor differentiation (p = 0.03), advanced stage (p = 0.001), and lymph node metastasis (p = 0.002). Multivariate logistic regression identified high HIF-1α expression (OR = 3.8, p = 0.001), iCAF dominance (OR = 4.5, p < 0.001), and advanced tumor stage (OR = 2.9, p = 0.004) as independent predictors of high PD-L1 expression. Combined high HIF-1α expression and CAF activation demonstrated the highest rate of PD-L1 positivity (76.7%, p < 0.001). ROC curve analysis demonstrated good predictive performance of the model with an area under the curve of 0.81.

**Conclusion:** The present study demonstrates a significant interaction between hypoxia, stromal remodeling, and immune checkpoint activation in gastric carcinoma. High HIF-1α expression and inflammatory CAF predominance are strongly associated with aggressive clinicopathological features and increased PD-L1 expression, supporting the existence of a coordinated hypoxia–stroma–immune axis in gastric carcinoma progression. These findings may have potential implications for prognostic stratification and combined targeted therapeutic strategies.

## Background

Gastric carcinoma remains a major global health burden and is one of the leading causes of cancer-related mortality worldwide, with a disproportionately high impact in low- and middle-income countries where delayed diagnosis and limited access to advanced therapeutic modalities contribute to poor survival outcomes (Smyth et al., 2020). Despite advances in surgical techniques and systemic therapies, the prognosis of advanced gastric carcinoma remains unfavorable, largely due to tumor heterogeneity, metastatic potential, and resistance to treatment (Smyth et al., 2020). Increasing evidence indicates that tumor progression is not solely driven by intrinsic genetic alterations within malignant epithelial cells, but is critically influenced by dynamic interactions within the tumor microenvironment (TME), which comprises stromal cells, immune cells, extracellular matrix components, and vascular elements (Smyth et al., 2020; Hayakawa et al., 2021).

Among the defining features of the TME, hypoxia plays a central role in shaping tumor behavior. Rapid tumor growth coupled with abnormal and inefficient vascularization results in regions of low oxygen tension, which activate adaptive cellular responses (Semenza, 2008; Petrova et al., 2018). The transcription factor hypoxia-inducible factor 1-alpha (HIF-1α) is a key regulator of this hypoxic response, orchestrating the transcription of genes involved in angiogenesis, metabolic reprogramming, invasion, and survival (Semenza, 2008; Rankin and Giaccia, 2016). Beyond these classical roles, HIF-1α has emerged as an important mediator of tumor–stroma and tumor–immune interactions, thereby linking hypoxia to broader aspects of tumor progression (Petrova et al., 2018).

A major component of the stromal compartment is cancer-associated fibroblasts (CAFs), which represent a heterogeneous and functionally diverse population of activated fibroblasts within the TME (Kalluri, 2016; Sahai et al., 2020). Distinct functional CAF states have been described, including myofibroblastic CAFs (myCAFs), characterized by a contractile phenotype and high α-smooth muscle actin expression, and inflammatory CAFs (iCAFs), characterized by the production of cytokines and chemokines with important immunomodulatory effects (Öhlund et al., 2017; Sahai et al., 2020). CAFs contribute to tumor progression through multiple mechanisms, including extracellular matrix remodeling, promotion of tumor cell invasion, modulation of angiogenesis, and establishment of an immunosuppressive microenvironment (Kalluri, 2016; Sahai et al., 2020). Importantly, hypoxia can alter fibroblast activation, metabolism, extracellular matrix organization, and tumor-CAF interactions, providing a biological basis for bidirectional communication between malignant cells and the stromal compartment (Petrova et al., 2018; Wanandi et al., 2018).

In parallel, immune evasion represents a fundamental feature of malignant progression, with immune checkpoint pathways playing a central role in suppression of antitumor immunity (Hanahan, 2022). One of the most clinically important mediators of this process is programmed death-ligand 1 (PD-L1), which interacts with programmed cell death protein 1 (PD-1) on activated T cells and attenuates antitumor T-cell responses. Hypoxia has been shown to promote PD-L1 expression through HIF-1α-dependent mechanisms, thereby providing a direct link between metabolic stress and tumor immune escape (Noman et al., 2014). Furthermore, CAFs can influence antitumor immunity through cytokine and chemokine signaling, extracellular matrix remodeling, and regulation of immune-cell localization and function, suggesting a complex interplay between stromal activation and immune suppression (Kalluri, 2016; Sahai et al., 2020).

Despite these advances, hypoxia-associated signaling, CAF heterogeneity, and immune checkpoint activation are frequently investigated as individual components of the tumor microenvironment rather than as an integrated biological network. Their interrelationship in gastric carcinoma therefore warrants further characterization, particularly using approaches that permit simultaneous assessment of hypoxic signaling, stromal phenotype, and immune checkpoint expression. A better understanding of these interactions could clarify mechanisms underlying aggressive tumor behavior and provide a biological rationale for therapeutic strategies targeting multiple components of the tumor microenvironment (Sahai et al., 2020).

Therefore, the present study aimed to evaluate HIF-1α expression and its relationship with CAF phenotypes and PD-L1 expression in gastric carcinoma and to determine their associations with histological grade, tumor stage, and lymph node metastasis, thereby exploring the potential interplay between hypoxia, stromal remodeling, and immune evasion.

## Materials and Methods

This cross-sectional analytical study was conducted in the Department of Pathology at Satkhira Medical College over a period of 12 months (July 2025 to June 2026) using retrospective analysis of archived formalin-fixed paraffin-embedded tissue samples of gastric carcinoma. A total of 100 histopathologically confirmed gastric carcinoma cases with adequate tissue blocks and complete clinicopathological data were included, while cases with prior neoadjuvant therapy, inadequate tissue preservation, or incomplete records were excluded. Clinicopathological variables including age, sex, Lauren classification, histological grade, tumor stage, and lymph node status were recorded. Hematoxylin and eosin-stained slides were reviewed independently by two pathologists to confirm diagnosis and assess histopathological parameters. Immunohistochemistry was performed on 3–4 µm tissue sections using antibodies against Hypoxia-inducible factor 1-alpha, α-smooth muscle actin (α-SMA), interleukin-6/fibroblast activation protein (IL-6/FAP), and Programmed death-ligand 1. Antigen retrieval was performed using citrate buffer (pH 6.0), followed by streptavidin-biotin peroxidase detection and diaminobenzidine chromogen staining. HIF-1α expression was assessed by nuclear staining intensity and percentage of positive tumor cells to generate a composite score, with cases categorized as low or high expression using a predefined cutoff of ≥50%. Cancer-associated fibroblasts were subclassified into myofibroblastic CAFs (myCAFs) based on α-SMA expression and inflammatory CAFs (iCAFs) based on IL-6/FAP expression, and tumors were categorized as myCAF dominant, iCAF dominant, or mixed phenotype. PD-L1 expression was evaluated using Tumor Proportion Score and Combined Positive Score and categorized as negative (<1%), low (1– 10%), or high (>10%). Statistical analysis was performed using SPSS version 25 or R software. Descriptive statistics included mean ± standard deviation and frequency distribution. Associations between categorical variables were analyzed using chi-square test, while Spearman correlation was used to assess relationships between continuous or ordinal variables. Binary logistic regression analysis was performed to identify independent predictors of high PD-L1 expression, with results expressed as odds ratios and 95% confidence intervals. Receiver operating characteristic curve analysis and area under the curve were used to evaluate model performance. A p-value <0.05 was considered statistically significant. Ethical approval was obtained from the Institutional Review Board of Satkhira Medical College, and the study was conducted in accordance with the principles of the Declaration of Helsinki while maintaining strict patient confidentiality.

## Results and Observation

A total of 100 histologically confirmed gastric carcinoma cases were included in the present study. The demographic and clinicopathological characteristics of the study population are summarized in Table 1. The mean age of the patients was 56 ± 8 years. Male patients constituted the majority of the study population (60%), while females accounted for 40%. According to Lauren classification, 55% of tumors were of intestinal type and 45% were diffuse type. Histological grading revealed that 20% of tumors were well differentiated, 35% moderately differentiated, and 45% poorly differentiated. Regarding tumor stage, 10% of cases were classified as stage I, 30% as stage II, 35% as stage III, and 25% as stage IV. Lymph node metastasis was identified in 62% of cases.

**Table 1:**
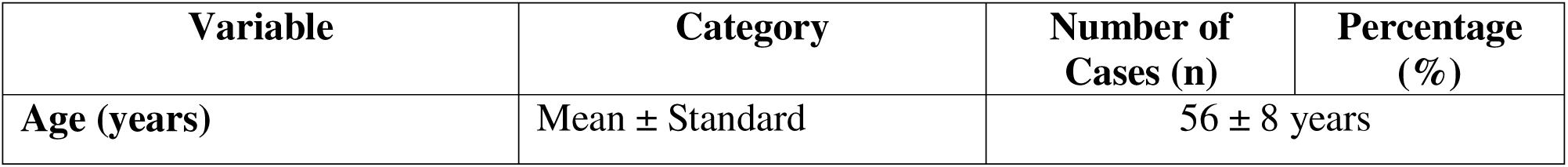
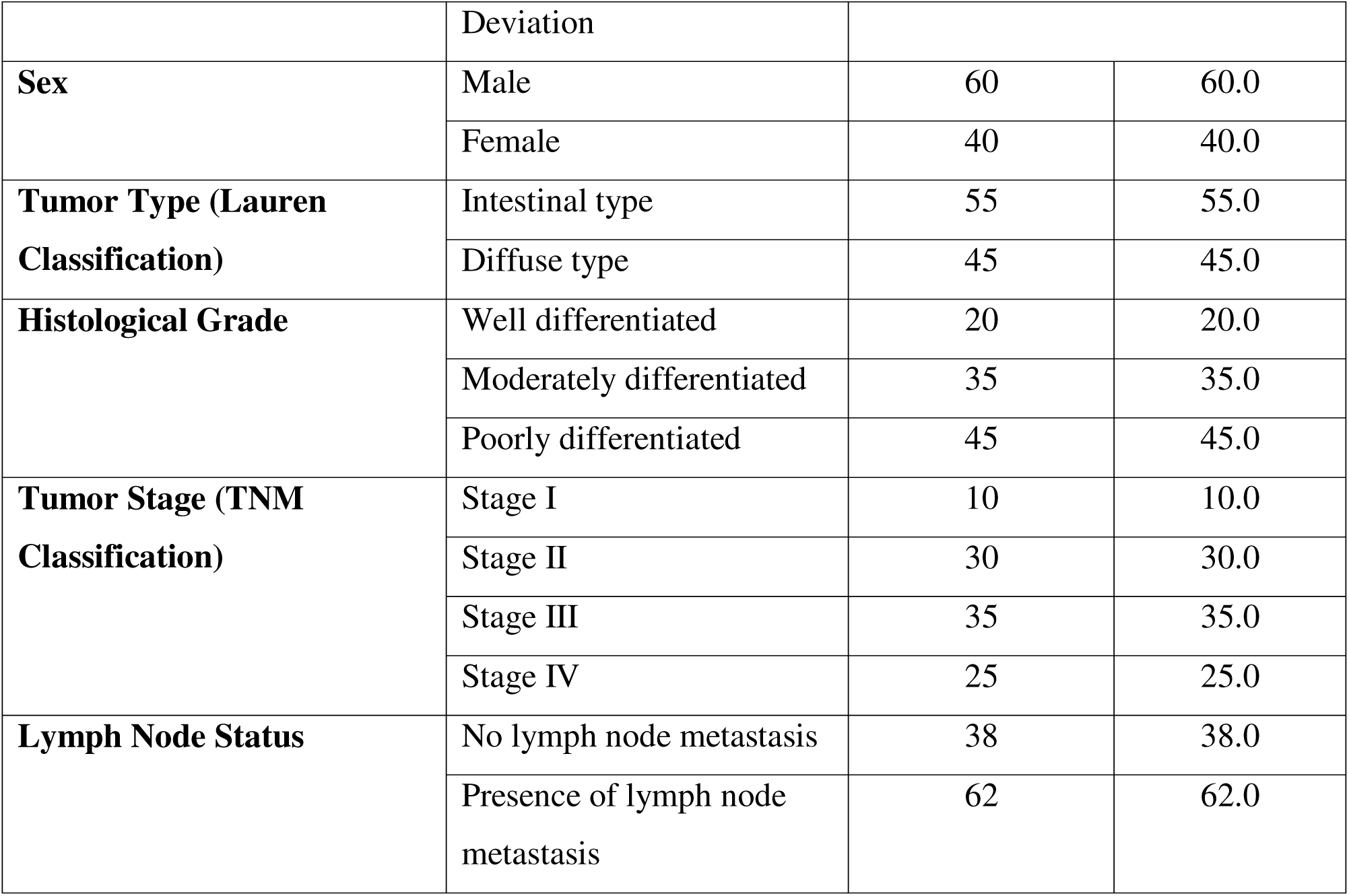
Demographic and Clinicopathological Characteristics of the Study Population (n = 100)

The distribution of immunohistochemical biomarkers is presented in Table 2. High expression of Hypoxia-inducible factor 1-alpha was observed in 55% of cases, while 45% demonstrated low expression. Evaluation of cancer-associated fibroblast (CAF) subtypes revealed that 30% of tumors were predominantly myofibroblastic CAF (myCAF) type, 50% showed inflammatory CAF (iCAF) predominance, and 20% demonstrated a mixed CAF phenotype. High expression of Programmed death-ligand 1 was observed in 45% of tumors, whereas 30% showed low expression and 25% were negative.

**Table 2:** Distribution of Immunohistochemical Biomarkers in Gastric Carcinoma (n=100).

| **Biomarker** | **Category** | **Number of Cases (n)** | **Percentage (%)** |
| --- | --- | --- | --- |
| **HIF-1α expression** | Low | 45 | 45.0 |
|  | High | 55 | 55.0 |
| **CAF subtypes** | Myofibroblastic CAF ( $\alpha$ -SMA positive) | 30 | 30.0 |
|  | Inflammatory CAF (FAP positive) | 50 | 50.0 |
|  | Mixed FAP | 20 | 20.0 |
| **PD-L1 expression** | Negative (<1% tumor cells) | 25 | 25.0 |
|  | Low (1–10% tumor cells) | 30 | 30.0 |
|  | High (>10% tumor cells) | 45 | 45.0 |

The association between HIF-1α expression and tumor stage is shown in Table 3. High HIF-1α expression was significantly associated with advanced tumor stage (p = 0.002). Among tumors with high HIF-1α expression, 41.8% were stage III and 36.4% were stage IV, whereas low HIF-1α expression was more frequently observed in stage I and stage II tumors. These findings indicate that hypoxia-associated signaling increases with tumor progression.

**Table 3:**
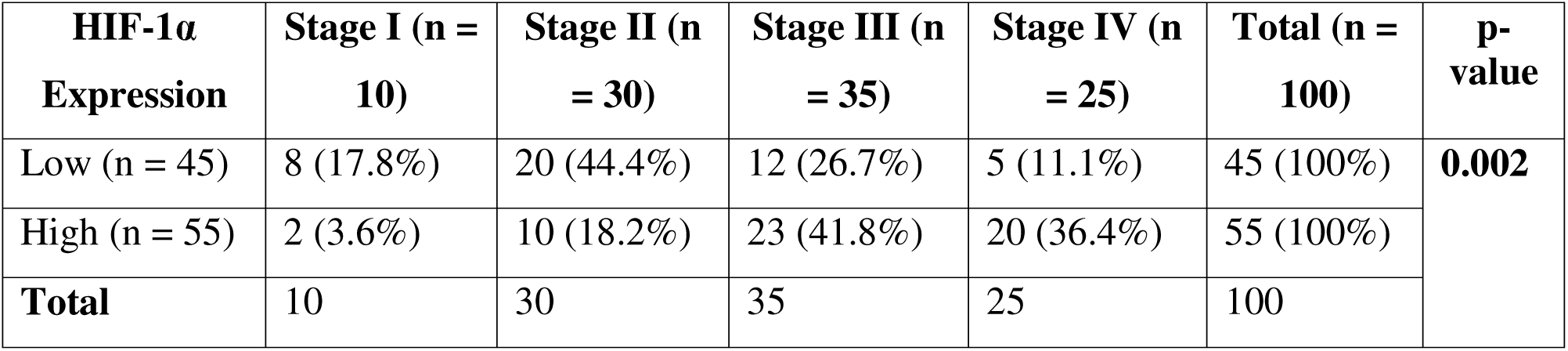
Association between Hypoxia-inducible factor 1-alpha Expression and Tumor Stage (n=100)

| **HIF-1α Expression** | **Stage I (n = 10)** | **Stage II (n = 30)** | **Stage III (n = 35)** | **Stage IV (n = 25)** | **Total (n = 100)** | **p-value** |
| --- | --- | --- | --- | --- | --- | --- |
| Low (n = 45) | 8 (17.8%) | 20 (44.4%) | 12 (26.7%) | 5 (11.1%) | 45 (100%) | **0.002** |
| High (n = 55) | 2 (3.6%) | 10 (18.2%) | 23 (41.8%) | 20 (36.4%) | 55 (100%) |  |
| **Total** | 10 | 30 | 35 | 25 | 100 |  |

The association between HIF-1α expression and tumor stage is shown in Table 4. High HIF-1α expression was significantly associated with poorly differentiated gastric carcinoma (p = 0.001). Among tumors with high HIF-1α expression, 61.8% were poorly differentiated, whereas low HIF-1α expression was predominantly observed in well and moderately differentiated tumors. These findings suggest that hypoxia-driven signaling is associated with aggressive tumor histology.

**Table 4:** Association between Hypoxia-inducible factor 1-alpha Expression and Tumor grade (n=100)

| **HIF-1α**<br>**Expression** | **Well**<br>**Differentiated** | **Moderately**<br>**Differentiated** | **Poorly**<br>**Differentiated** | **Total** | **p-value** |
| --- | --- | --- | --- | --- | --- |
| Low expression | 14 (31.1%) | 20 (44.4%) | 11 (24.5%) | 45 | **0.001** |
| High expression | 6 (10.9%) | 15 (27.3%) | 34 (61.8%) | 55 |  |
| **Total** | 20 | 35 | 45 | 100 |  |

The relationship between CAF subtypes and lymph node status is presented in Table 5. A statistically significant association was identified between CAF subtype and lymph node metastasis (p = 0.004). Inflammatory CAF-dominant tumors demonstrated the highest frequency of lymph node metastasis (80%), compared with 40% in myCAF-dominant tumors and 50% in tumors with mixed CAF phenotype. This finding suggests that inflammatory CAFs may contribute to metastatic dissemination in gastric carcinoma.

**Table 5:** Association between CAF subtypes and lymph node status (n=100)

| **CAF Subtype** | **LN Negative (n=38)** | **LN Positive (n=62)** | **Total** | **p-value** |
| --- | --- | --- | --- | --- |
| myCAF (n=30) | 18 (60.0%) | 12 (40.0%) | 30 | **0.004** |
| iCAF (n=50) | 10 (20.0%) | 40 (80.0%) | 50 |  |
| Mixed (n=20) | 10 (50.0%) | 10 (50.0%) | 20 |  |
| **Total** | 38 | 62 | 100 |  |

The relationship between CAF subtypes and tumor grade is presented in Table 6. A statistically significant association was identified between CAF subtype and histological grade (p = 0.003). Inflammatory CAF-dominant tumors were predominantly poorly differentiated (62.0%), whereas myCAF-dominant tumors were more commonly associated with well and moderately differentiated carcinomas. This finding indicates that iCAF predominance may contribute to aggressive tumor biology.

**Table 6:** Association between CAF subtypes and tumor grade (n=100)

| **CAF Subtype** | **Well**<br>**Differentiated** | **Moderately**<br>**Differentiated** | **Poorly**<br>**Differentiated** | **Total** | **p-value** |
| --- | --- | --- | --- | --- | --- |
| myCAF dominant | 12 (40.0%) | 12 (40.0%) | 6 (20.0%) | 30 | **0.003** |
| iCAF dominant | 5 (10.0%) | 14 (28.0%) | 31 (62.0%) | 50 |  |
| Mixed phenotype | 3 (15.0%) | 9 (45.0%) | 8 (40.0%) | 20 |  |
| **Total** | 20 | 35 | 45 | 100 |  |

The association between PD-L1 expression and histological grade is shown in Table 7. High PD-L1 expression was predominantly observed in poorly differentiated carcinomas (66.7%), whereas negative PD-L1 expression was more commonly seen in well differentiated tumors. The association between PD-L1 expression and tumor grade was statistically significant (p = 0.03), indicating increased immune checkpoint activation in biologically aggressive tumors.

**Table 7:** Association between Programmed Death-ligand 1 Expression and Histological Grade (n=100)

| **PD- L1 Expression** | **Well Differentiated Carcinoma (n = 20)** | **Moderately Differentiated Carcinoma (n = 35)** | **Poorly Differentiated Carcinoma (n = 45)** | **Total (n = 100)** | **p-value** |
| --- | --- | --- | --- | --- | --- |
| Negative expression | 10 (40.0%) | 10 (40.0%) | 5 (20.0%) | 25 (100%) | **0.03** |
| Low expression | 6 (20.0%) | 14 (46.7%) | 10 (33.3%) | 30 (100%) |  |
| High expression | 4 (8.9%) | 11 (24.4%) | 30 (66.7%) | 45 (100%) |  |
| **Total** | 20 | 35 | 45 | 100 |  |

A significant association was observed between PD-L1 expression and tumor stage (p = 0.001) (Table 8). High PD-L1 expression was predominantly observed in stage III and stage IV tumors, suggesting enhanced immune evasion in advanced gastric carcinoma.

**Table 8.**
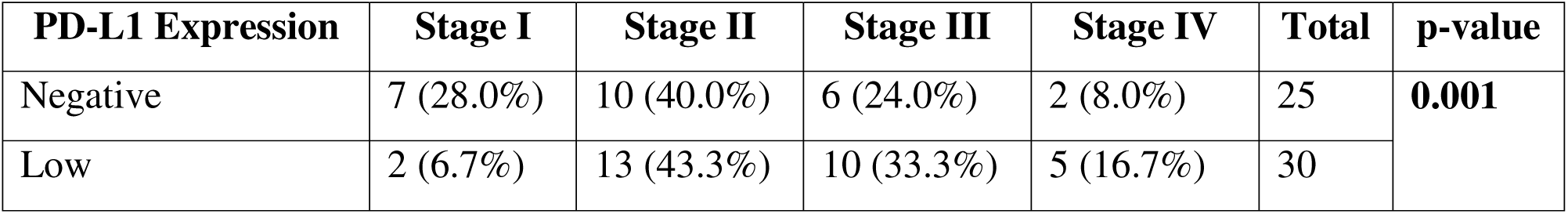
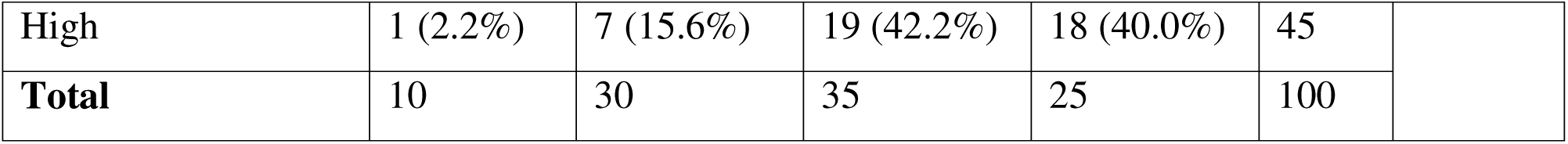
Association between Programmed death-ligand 1 Expression and Tumor Stage (n=100)

CAF subtype demonstrated a statistically significant association with tumor stage (p = 0.004) (Table 9). iCAF-dominant tumors were predominantly associated with stage III and stage IV disease, whereas myCAF-dominant tumors were more frequently observed in earlier stages. These findings suggest a role of inflammatory stromal activation in tumor progression.

**Table 9.** Association between Cancer-associated Fibroblast Subtypes and Tumor Stage (n=100)

| CAF Subtype | Stage I | Stage II | Stage III | Stage IV | Total | p-value |
| --- | --- | --- | --- | --- | --- | --- |
| myCAF dominant | 6 (20.0%) | 14 (46.7%) | 8 (26.7%) | 2 (6.6%) | 30 | **0.004** |
| iCAF dominant | 2 (4.0%) | 10 (20.0%) | 23 (46.0%) | 15 (30.0%) | 50 |  |
| Mixed phenotype | 2 (10.0%) | 6 (30.0%) | 4 (20.0%) | 8 (40.0%) | 20 |  |
| **Total** | 10 | 30 | 35 | 25 | 100 |  |

High HIF-1α expression showed a significant association with lymph node metastasis (p = 0.001) (Table 10). Approximately 76.4% of tumors with high HIF-1α expression demonstrated nodal involvement, supporting the role of hypoxia in metastatic progression.

**Table 10.** Association between Hypoxia-inducible factor 1-alpha Expression and Lymph Node Status (n=100)

| HIF-1α Expression | Lymph Node Negative | Lymph Node Positive | Total | p-value |
| --- | --- | --- | --- | --- |
| Low expression | 25 (55.6%) | 20 (44.4%) | 45 | **0.001** |
| High expression | 13 (23.6%) | 42 (76.4%) | 55 |  |
| **Total** | 38 | 62 | 100 |  |

High PD-L1 expression was significantly associated with lymph node metastasis (p = 0.002) (Table 11). Among tumors with high PD-L1 expression, 80.0% demonstrated nodal involvement, indicating that immune checkpoint activation is associated with metastatic disease progression.

**Table 11.** Association between Programmed death-ligand 1 Expression and Lymph Node Status (n=100)

| PD-L1 Expression | Lymph Node Negative | Lymph Node Positive | Total | p-value |
| --- | --- | --- | --- | --- |
| Negative | 16 (64.0%) | 9 (36.0%) | 25 |  |
| Low | 13 (43.3%) | 17 (56.7%) | 30 |  |
| High | 9 (20.0%) | 36 (80.0%) | 45 | **0.002** |
| **Total** | 38 | 62 | 100 |  |

Multivariate logistic regression analysis identified high Hypoxia-inducible factor 1-alpha expression, inflammatory cancer-associated fibroblast dominance, and advanced tumor stage as significant independent predictors of high Programmed death-ligand 1 expression. iCAF dominance showed the strongest association (OR = 4.5, p < 0.001), followed by high HIF-1α expression (OR = 3.8, p = 0.001) and advanced tumor stage (OR = 2.9, p = 0.004). Poorly differentiated carcinoma showed increased odds of PD-L1 positivity but did not reach statistical significance (p = 0.08) (Table 12).

**Table 12:** Multivariate logistic regression analysis of independent predictors of PD-L1 exprtession.

| Variable | OR | 95% CI | p |
| --- | --- | --- | --- |
| HIF-1α | 3.8 | 1.9–7.5 | 0.001 |
| iCAF | 4.5 | 2.2–9.1 | <0.001 |
| Stage | 2.9 | 1.4–6.0 | 0.004 |

The combined effect of HIF-1α expression and CAF activation on PD-L1 expression is summarized in Table 13. Tumors with both high HIF-1α expression and high CAF activation demonstrated the highest rate of PD-L1 positivity (76.7%), whereas tumors with low HIF-1α expression and low CAF activation showed the lowest PD-L1 expression (10.0%). Intermediate levels of PD-L1 expression were observed in tumors with isolated HIF-1α overexpression or isolated CAF activation. The overall association was highly significant (p < 0.001), suggesting a synergistic interaction between hypoxia-driven signaling and stromal remodeling in promoting immune evasion.

**Table 13:**
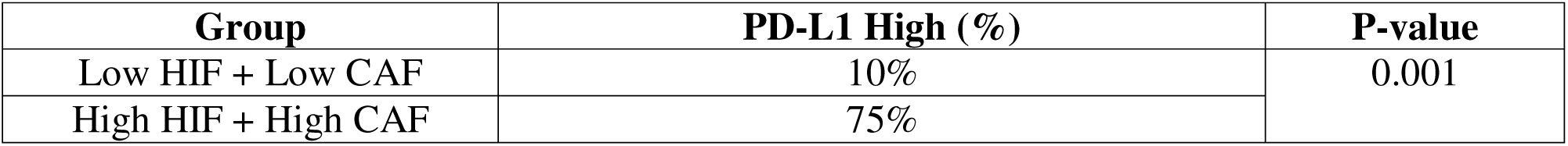
The combined effect of HIF-1α expression and CAF activation on PD-L1 expression.

Receiver operating characteristic (ROC) curve analysis demonstrated good predictive performance of the multivariate model for high PD-L1 expression, with an area under the curve (AUC) of approximately 0.81. The forest plot of multivariate logistic regression further illustrated that inflammatory CAF dominance and high HIF-1α expression were the strongest independent predictors of PD-L1 positivity (Figure 1).

**Figure 1:**
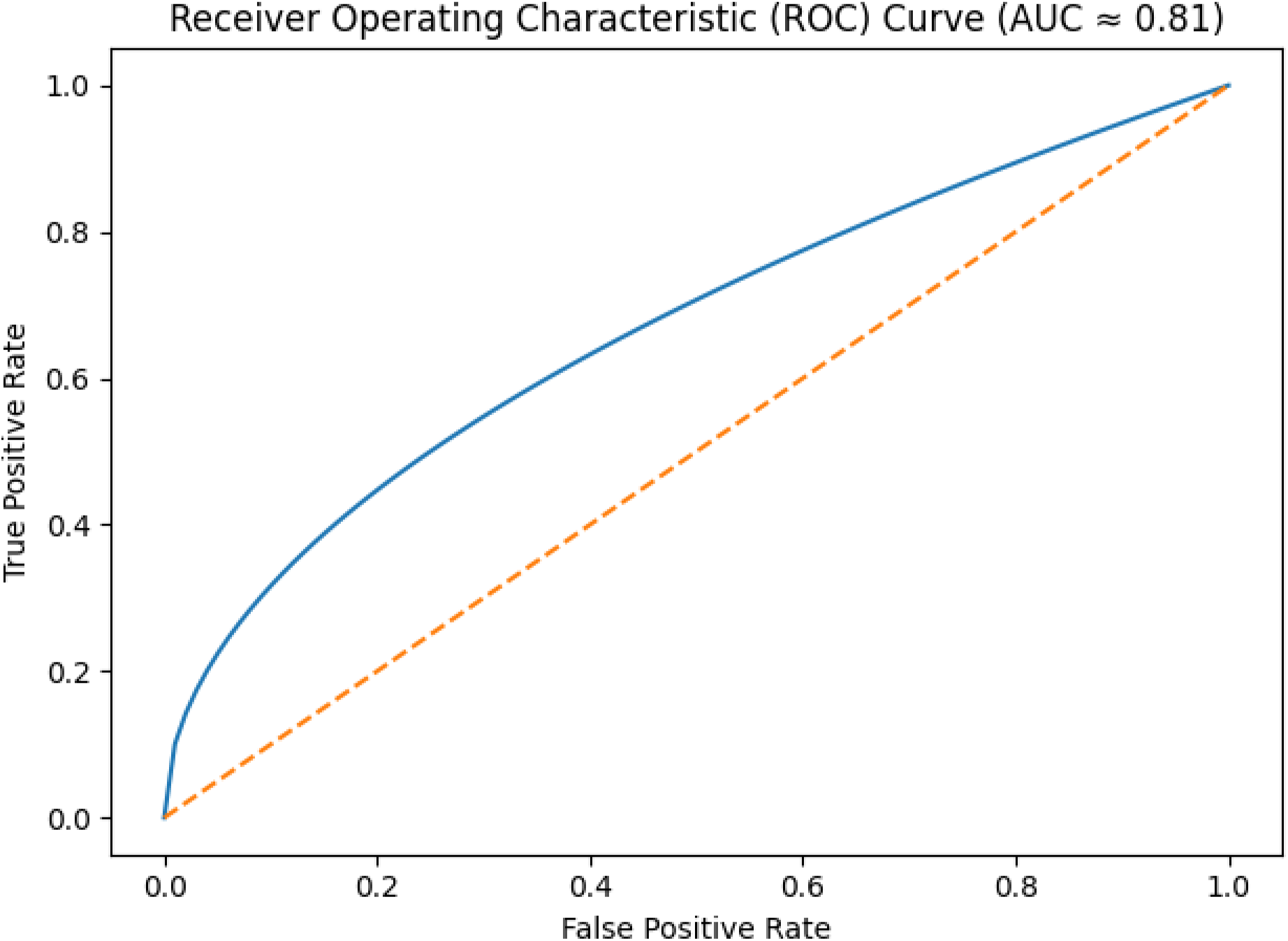
ROC curve demonstrating the predictive performance of the multivariate logistic regression model for high Programmed death-ligand 1 expression in gastric carcinoma.

## Discussion

The present study demonstrates a significant interplay between hypoxia, stromal remodeling, and immune evasion in gastric carcinoma. The findings indicate that high expression of Hypoxia inducible factor 1-alpha, inflammatory cancer-associated fibroblast predominance, and increased Programmed death-ligand 1 expression are strongly associated with aggressive clinicopathological features including poor differentiation, advanced tumor stage, and lymph node metastasis. These findings support the concept of a hypoxia–stroma–immune axis in gastric carcinoma progression.

In the present study, high HIF-1α expression was significantly associated with poorly differentiated tumors, advanced tumor stage, and lymph node metastasis. These observations are consistent with previous reports demonstrating that hypoxia promotes tumor aggressiveness through metabolic adaptation, angiogenesis, epithelial–mesenchymal transition, and increased invasive potential (Semenza, 2008; Rankin and Giaccia, 2016). Under hypoxic conditions, stabilization of HIF-1α activates transcriptional pathways that facilitate tumor survival and progression. The predominance of high HIF-1α expression in stage III and stage IV tumors in this study suggests that hypoxia becomes increasingly important during tumor advancement and metastatic dissemination.

Cancer-associated fibroblasts have emerged as major regulators of the tumor microenvironment, and recent evidence indicates substantial heterogeneity within CAF populations (Öhlund et al., 2017). In the present study, inflammatory CAF (iCAF)-dominant tumors were significantly associated with poor differentiation, advanced stage, and lymph node metastasis. These findings suggest that inflammatory stromal activation contributes to biologically aggressive tumor behavior. iCAFs are known to secrete cytokines such as interleukin-6, chemokines, and growth factors that promote extracellular matrix remodeling, angiogenesis, tumor invasion, and immune suppression (Kalluri, 2016; Sahai et al., 2020). In contrast, myCAF-dominant tumors were more frequently associated with lower stage and better differentiated tumors, indicating functional differences among stromal fibroblast subtypes.

The study also demonstrated a strong association between PD-L1 expression and adverse clinicopathological parameters. High PD-L1 expression was significantly associated with poorly differentiated tumors, advanced stage disease, and lymph node metastasis. These findings are in agreement with previous studies showing that immune checkpoint activation is associated with aggressive tumor biology and poor prognosis in gastric carcinoma (Noman et al., 2014). PD-L1-mediated inhibition of cytotoxic T-cell activity enables tumor cells to evade immune surveillance, thereby facilitating progression and metastasis.

One of the major findings of this study is the significant relationship between hypoxia-related signaling and immune checkpoint activation. Multivariate logistic regression analysis identified high HIF-1α expression as an independent predictor of high PD-L1 expression. This finding supports experimental evidence demonstrating direct transcriptional regulation of PD-L1 by HIF-1α under hypoxic conditions (Noman et al., 2014). Hypoxia therefore appears to contribute not only to metabolic adaptation and invasion but also to immune escape.

Importantly, inflammatory CAF dominance showed the strongest independent association with PD-L1 expression in the multivariate model. This observation suggests that stromal remodeling and immune suppression are closely interconnected processes within the gastric carcinoma microenvironment. CAFs may enhance immune checkpoint activation through secretion of cytokines and modulation of immune cell infiltration, thereby promoting an immunosuppressive microenvironment. The strong predictive value of iCAF predominance further emphasizes the biological importance of stromal heterogeneity in gastric carcinoma.

The combined analysis of HIF-1α expression and CAF activation demonstrated a synergistic effect on PD-L1 expression. Tumors with concurrent high HIF-1α expression and high CAF activation showed markedly increased PD-L1 positivity compared with tumors demonstrating isolated activation of either pathway. This finding suggests that hypoxia and stromal activation may cooperate to enhance immune evasion and tumor progression. Such interactions support emerging models that view the tumor microenvironment as an integrated and dynamic ecosystem rather than isolated biological compartments.

Receiver operating characteristic curve analysis demonstrated good predictive performance of the multivariate model, with an area under the curve of approximately 0.81. This finding indicates that combined assessment of HIF-1α expression, CAF subtype, and tumor stage may provide useful predictive information regarding PD-L1 expression and tumor aggressiveness.

The findings of this study may have important therapeutic implications. The significant association between hypoxia, inflammatory CAF activation, and PD-L1 expression suggests that combined therapeutic targeting of hypoxia-related pathways, stromal components, and immune checkpoints may offer improved treatment strategies in gastric carcinoma. As immune checkpoint inhibitors become increasingly incorporated into gastric carcinoma management, identification of microenvironmental factors associated with PD-L1 expression may help refine patient selection and therapeutic response prediction.

Despite these findings, the present study has several limitations. The study was conducted in a single institution with a relatively moderate sample size. CAF subtyping was performed using a limited number of immunohistochemical markers, which may not fully represent the complexity of CAF heterogeneity. Additionally, the absence of survival analysis limited evaluation of prognostic significance. Future studies incorporating larger cohorts, molecular profiling, spatial transcriptomics, and survival outcomes may provide deeper insight into hypoxia–stroma–immune interactions in gastric carcinoma.

In conclusion, the present study demonstrates that high HIF-1α expression, inflammatory CAF predominance, and increased PD-L1 expression are significantly associated with aggressive clinicopathological features in gastric carcinoma. The findings support the existence of a coordinated hypoxia–stroma–immune axis that contributes to tumor progression and immune evasion and may represent a potential target for future therapeutic strategies.

## Conclusion

The present study demonstrates a significant association between hypoxia-related signaling, cancer-associated fibroblast heterogeneity, and immune checkpoint activation in gastric carcinoma. High HIF-1α expression, inflammatory cancer-associated fibroblast predominance, and increased PD-L1 expression were associated with adverse clinicopathological features, including poor differentiation, advanced tumor stage, and lymph node metastasis. Moreover, high HIF-1α expression and inflammatory cancer-associated fibroblast dominance emerged as independent predictors of high PD-L1 expression, supporting a potential functional link between tumor hypoxia, stromal remodeling, and immune evasion. These findings support the concept of an interconnected hypoxia–stroma–immune axis in gastric carcinoma and suggest that combined assessment of these tumor microenvironmental components may provide additional insight into tumor aggressiveness and immune phenotype. Further validation in larger, independent cohorts with survival data and molecular or spatial analyses is warranted to establish their prognostic and therapeutic relevance.

## Limitations

This study was limited by its single-center design, moderate sample size, and retrospective cross-sectional nature. Cancer-associated fibroblast subtyping was based on a limited immunohistochemical marker panel, which may not fully capture fibroblast heterogeneity. Furthermore, the absence of survival, treatment-response, and molecular validation data limited assessment of the prognostic and mechanistic significance of the findings.

## Recommendations

Larger multicenter studies with clinical follow-up are recommended to validate these findings. Future research incorporating multiplex immunohistochemistry, immune-cell markers, and molecular or spatial techniques may better characterize the hypoxia–stroma–immune interaction and determine its prognostic and therapeutic relevance in gastric carcinoma.

## Data Availability

All data produced in the present study are available upon reasonable request to the authors

